# Geospatial-based neighborhood hazards and asthma risk in adults: interaction with social vulnerability

**DOI:** 10.1101/2025.06.29.25330503

**Authors:** Yujing Chen, Ming Kei Chung

## Abstract

**Background:** Various residential hazards have been associated with asthma risk, with socially disadvantaged neighborhoods disproportionately affected by environmental exposures. This study aimed to explore geospatial-based neighborhood hazards and estimate their mixture effect on adult asthma risk by social vulnerability index (SVI).

**Materials and Methods:** This cross-sectional study included 7092 adults from a North Carolina-based cohort (2013-2022). Adult asthma was defined as self-reported an asthma diagnosis with recurrent attacks in adulthood. Exposure to 29 residential hazards was estimated using spatiotemporal models and hazard maps, including multiple air pollutants, volatile organic compound emissions, highway density, and proximity to pollutant sources. The census tract-based SVI was assigned to participants’ residences. Associations between specific hazards and asthma were examined using logistic regression in high and low SVI groups (stratified by median percentile), and joint effects of mixture exposure were analyzed using quantile g-computation.

**Results:** Residential proximity to dry-cleaning solvent contamination was positively associated with asthma in the overall population. In the high SVI group, seven additional hazards—particularly air pollutants—were significantly linked to increased asthma odds (False discovery rate, FDR< 0.05). A quintile increase in the mixture exposure was associated with a 35% (95% CI: 6%, 73%) higher odds of asthma in the high SVI group, which was more pronounced than in the low SVI group (*P* _for difference_= 0.038). Key contributors included toluene, dry-cleaning sites, organic matter in PM_2.5_, and carbon monoxide, each accounting for over 10% of the mixture effect. Socioeconomic and housing/transportation-related vulnerabilities interacted with key hazards and mixture exposure on asthma (FDR _interaction_ < 0.05).

**Conclusion:** Residents of high SVI neighborhoods are more vulnerable to environmental hazard-related asthma risks. Targeted interventions should prioritize communities with poor socioeconomic, housing, and transportation conditions to effectively mitigate these environmental health risks.

## Introduction

Asthma, a prevalent chronic respiratory disease, affects 262 million people worldwide and contributes significantly to morbidity and premature mortality in adults. ^1^ The onset and development of asthma are caused by complex interactions between host and environment, with neighborhood factors contributing through various adverse environmental and social exposures. Environmental hazards—primarily derived from anthropogenic sources (e.g., industry, agriculture, transportation, and energy production) ^2^— could pose cumulative health risks to individuals in their living environments. Research has recognized the important roles of residential hazards in asthma etiology ^3–5^. For example, higher residential traffic densities ^3^ and exposure to particulate matter and gaseous pollutants have been associated with declined lung function^6^ and increased asthma incidence. ^7^ Proximity to pollution sources, such as concentrated animal feeding operations or industrial facilities, has also been associated with increased risks of immune-related diseases and asthma. ^4^

However, previous epidemiological studies often relied on single-exposure approaches to explore asthma risk factors, potentially obscuring the health effect of hazard mixtures as a whole. Few recent studies have employed clustering methods to examine multi-exposure profiles in urban or other settings, exploring their associations with asthma. ^8–10^ For example, European cohort studies identified that exposome clusters characterized by high exposure to air pollutants, high built environment, and low greenness were associated with increased risk of adult or childhood asthma. ^8,10^ Nonetheless, these studies inadequately evaluated correlation patterns and mixture effects of neighborhood hazards from multiple toxicant sources. Another research gap persists in identifying socially vulnerable groups disproportionately affected by environmental exposures. Low-income and poor-quality housing communities often reside near highly polluted sources ^11,12^, where residents may experience high levels of pollution from multi-sources ^13^, driving an inequitable distribution of asthma risk. ^14,15^ Although socioeconomic characteristics and environmental hazards have been separately associated with asthma outcomes, few studies have contextualized exposures within disadvantaged neighborhoods. ^14,16,17^ Characterizing neighborhood factors driving vulnerability to hazard effects may help prioritize community-level interventions for asthma prevention.

Geospatial data have been integrated into the exposomic framework to infer individual exposure to environmental toxicants and relationship with health risk in populations. ^18^ The census-derived social vulnerability index (SVI), a composite measure reflecting community resilience, can also be spatially mapped to participants to quantify socioeconomic differences. ^2^ Utilizing geospatial data from a North-Carolina based cohort, this study estimated neighborhood environmental hazards and examined their individual and combined associations with adult asthma by social vulnerability groups. We hypothesized to identify key environmental hazards and their mixture effects on asthma through exposomic and mixture approaches. Domain-specific SVI components were examined to characterize vulnerable subpopulations that should be targeted for precision prevention and intervention of asthma.

## Methods

### Study design and population

The Personalized Environment and Genes Study (PEGS) was established in 2002 at the National Institute of Environmental Health Sciences (NIHES), which recruited demographically representative and diverse participants at over 200 unique sites in North Carolina. ^19^ From 2013 to 2022, 9449 adults (>18 years) participated in the cross-sectional Health and Exposure Survey, with 7770 provided completed residential addresses and asthma outcome information (version Freeze 3.1). This study further excluded participants with missing data in sociodemographic variables [i.e., age (n=91), race (n=205), and employment, education and income level (n=382)], resulting in 7092 PEGS participants in the final analysis. ^20^ The PEGS study protocol was approved by the NIEHS institutional review boards, and all participants provided written informed consent. The Survey and Behavioural Research Ethics Committee at the Chinese University of Hong Kong approved this study protocol (SBRE-23-0352).

### Geospatial exposure measurements

Geographic Information System (GIS) is a powerful computer mapping and analysis technology that allows large quantities of information to be analyzed within a geographic context. Participant’s residential address was geocoded into geographic coordinates and linked with hazardous exposure using GIS data. Twenty-nine GIS-based exposures encompass atmospheric composition (11 air pollutants), Toxics Release Inventory (TRI) [four volatile organic compounds (VOCs)], highway density (one variable), and proximity to hazardous point sources (13 pollutant sites). These exposures were estimated using established spatiotemporal model or hazard maps, and their data sources were further described in **Table 1**. This study calculated individual long-term average exposures by aggregating annual concentrations atmospheric composition across 2000-2018 or matching available most recent hazard maps during survey years. Exposures estimated within a 10-km buffer were prioritized for analysis due to the largest observed variations.

**Table 1.**
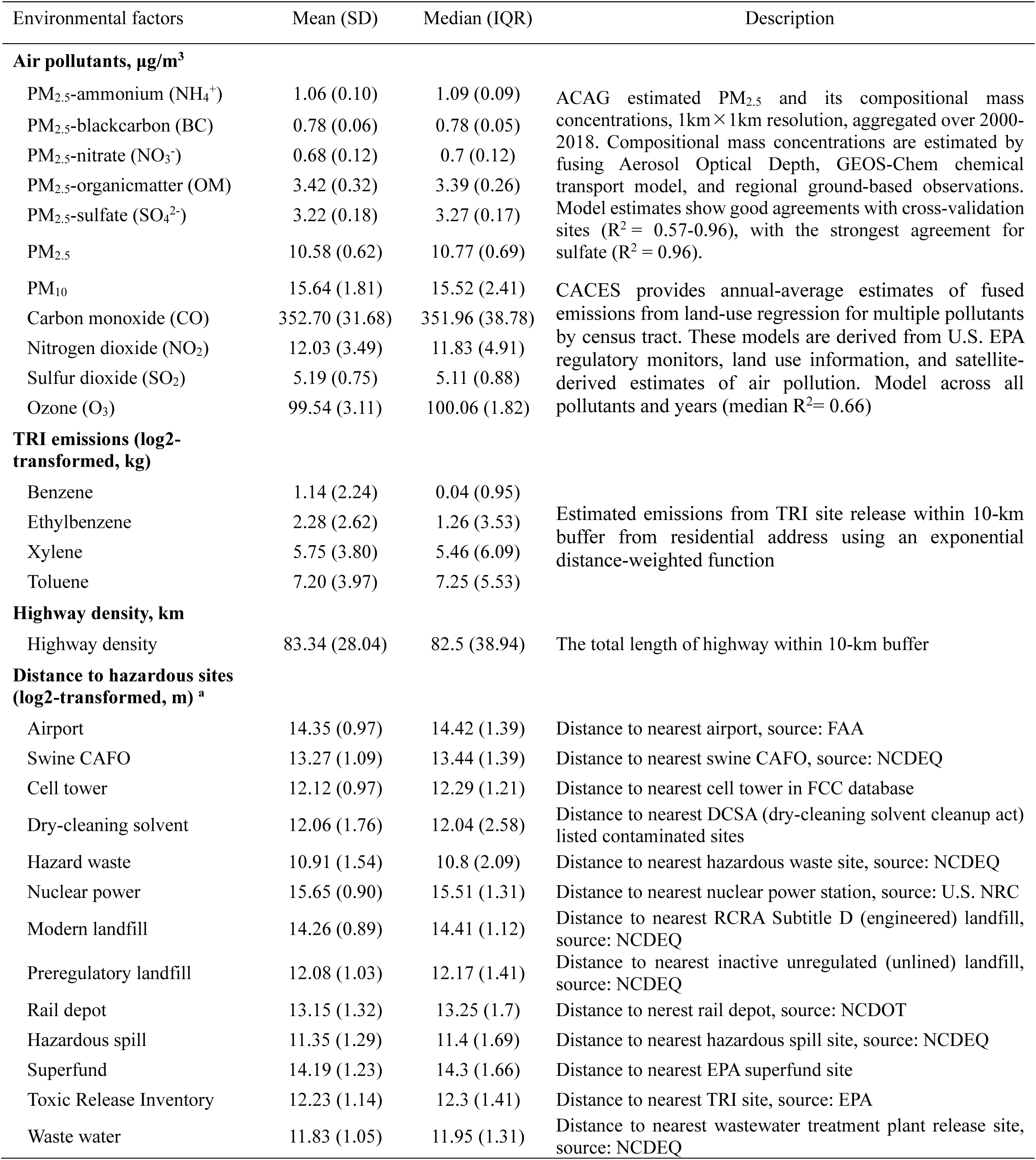
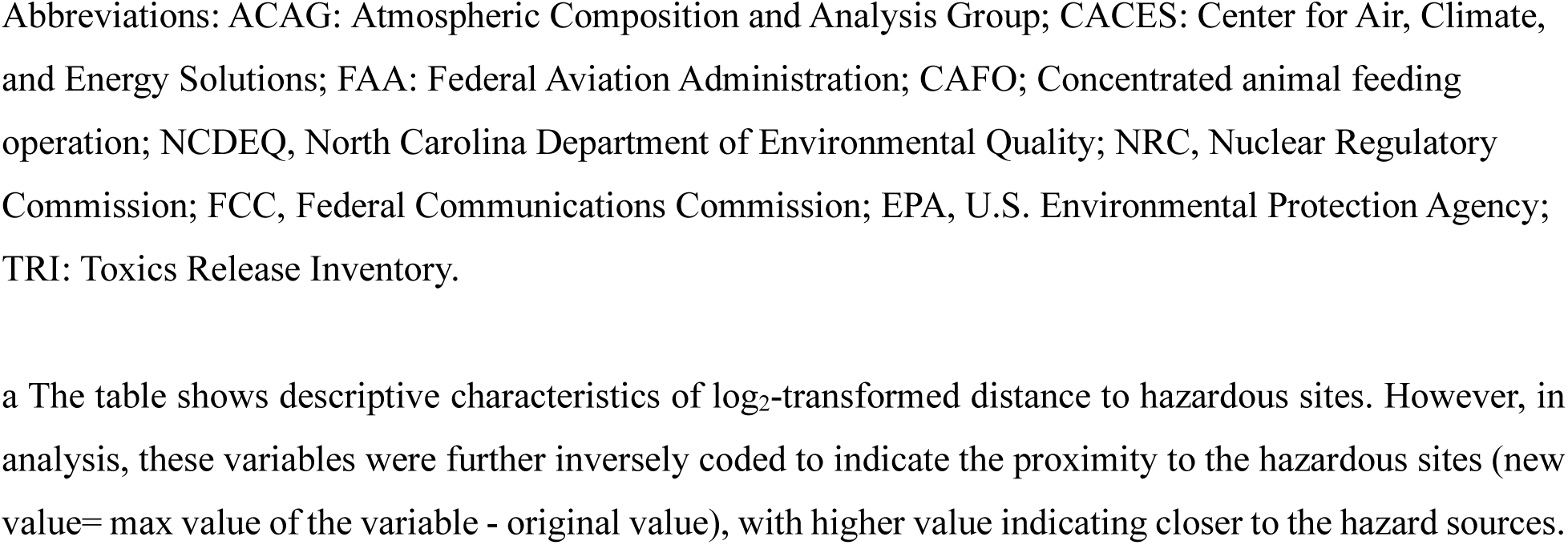
Description and distribution of hazard exposures surrounding participants’ residence.

### Social vulnerability index

The social vulnerability index (SVI) is a composite score designed by the Centers for Disease Control and Prevention (CDC) to consistently quantify neighborhood disadvantage across the United States over time ^21^. The index consists of an overall percentile ranking based on 15 demographic and socioeconomic factors that adversely affect communities facing hazards and other stressors. It includes four themes, namely socioeconomic status, household composition and disability, minority and language status, and housing and transportation type (see **Table S1**). We linked the census tract location of residence to the corresponding SVI for the closest reference years with available data (i.e., 2014, 2016, and 2018 for SVI).

### Asthma outcomes

Asthma diagnosis was determined by an affirmative answer to the question, “Has a doctor or other healthcare provider ever told you that you have asthma?” Active adult asthma was defined if individuals with asthma diagnosis reported that they still had asthma in adulthood or they experienced asthma attacks, emergency room visits, or received medication prescriptions in the past 12 months. PEGS survey questions were developed based on validated questionnaires in the Nurses’ Health Study II, National Health and Nutrition Examination Survey (NHANES), and the PhenX toolkit.^22^

### Covariates

Demographic characteristics were collected through self-administered questionnaires, including age, sex, race (White, Black, and others), highest education levels (high school or below, college, technical or vocational school graduate, bachelor, and graduate or professional), annual household income (less than $20,000, $20,000 to 49,999, $50,000 to 79,999, $80,000 or above), employment status (currently employed or not), smoking status (smoker>100 cigarettes in lifetime; never, former smoker, current smoker), and body mass index (BMI, kg/m^2^) calculated based on self-reported height and weight.

### Statistical analysis

General characteristics of participants were summarized as means and standard deviations (SDs) for continuous variables or frequencies and percentages for categorized variables. Differences between asthma and non-asthma groups were compared using student *t*-tests or χ^2^ tests. Exposures not following a normal distribution were log_2_-transformed, and their distribution was described in the total population and SVI subgroups. To enhance interpretations, the distance to hazardous sites was inversely coded to reflect proximity (new value = max value of the variable - original value) in the correlation and association analyses, with higher values indicating closer proximity to the hazard sources. Exposome correlation globes ^23^ were plotted to visualize pairwise correlations of environmental hazards by SVI.

Logistic regression was utilized to assess the pairwise association between each hazard and asthma with an exposome-wide association studies (ExWAS) approach.^24^ The main analytical model was adjusted for covariates selected from literature that are both associated with exposure and outcomes, including age, sex, race, employment, highest education levels, annual household income, smoking, and BMI. Multiple testing was corrected using Benjamini-Hochberg procedure false discovery rate (FDR). Additionally, ExWAS analysis was conducted for high SVI and low SVI groups, stratified by the median overall SVI. For exposures with FDR < 0.10 in the high overall SVI group, their interactions between specific SVI themes were examined by including the multiplicative term in the model.

The quantile g-computation model has been applied to assess mixture effects and identify key contributing hazards.^25,25,26^ The quantile g-computation method can address potential collinearity among exposures and allow the directional homogeneity assumption. ^27^ We applied this method and used quintiles to construct the quantile indicator variables representing the exposures. The weight indexes with effect direction were calculated for each hazard in the total population and in the SVI subgroup. We conducted two-sample z-tests to compare the mixture exposure effects between the lower and higher SVI groups, using stratum-specific estimates and standard errors in mixture models.

The potential non-linear hazard-asthma relationship for exposures with *p* < 0.05 in ExWAS was fitted by the restricted cubic splines (knots=3).We conducted several sensitivity analyses and visualized results using specification curves ^28^: (1) examining exposures estimated within different buffers (i.e., 5-km buffer for TRI emissions and highway density); (2) testing for the influence of parental asthma (self-reported diagnosis of asthma of parents) on the observed associations. Statistical analyses were conducted with R 4.5.0., using “*qgcomp*”, “rcssci”, “*Hmisc*” packages. A two-sided *P* value /*FDR* <0.05 was considered significant in multi-exposure analyses / multiple testing.

## Results

### General characteristics and asthma outcomes

As shown in **Table 2**, among the 7092 participants, the mean age is 50.97 (SD=15.44) years, with predominance of females (67.46%) and White individuals (73.21%). Current asthma was reported by 747 (10.53%) adults. Compared to non-asthma participants, asthma cases were younger, more likely to be female, Black, or smokers, and had lower education levels, lower household income, and higher BMI (*P*< 0.05). The asthma group also exhibited a higher percentile ranking in overall and theme-specific SVI, including socioeconomic, household composition, housing/transportation.

**Table 2.** General characteristics of study population (N=7092)

| Characteristics | Total<br>(N=7092) | Non-asthma<br>(n=6345) | Current-Asthma<br>(n=747) | *p* value |
| --- | --- | --- | --- | --- |
| **Individual sociodemographic characteristics** |  |  |  |  |
| Age, year, Mean (SD) | 50.97 (15.44) | 51.16 (15.56) | 49.38 (14.31) | 0.003 |
| Sex, n (%) |  |  |  | < 0.001 |
| Male | 2308 (32.54) | 2156 (33.98) | 152 (20.35) |  |
| Female | 4784 (67.46) | 4189 (66.02) | 595 (79.65) |  |
| Race, n (%) |  |  |  | 0.002 |
| White | 5192 (73.21) | 4682 (73.79) | 510 (68.27) |  |
| Black | 1625 (22.91) | 1416 (22.32) | 209 (27.98) |  |
| Others | 275 (3.88) | 247 (3.89) | 28 (3.75) |  |
| Highest education level, n (%) |  |  |  | < 0.001 |
| High school or below | 1178 (16.61) | 1024 (16.14) | 154 (20.62) |  |
| College, technical or vocational school graduate | 2191 (30.89) | 1939 (30.56) | 252 (33.73) |  |
| Bachelor | 1948 (27.47) | 1772 (27.93) | 176 (23.56) |  |
| Graduate or professional | 1775 (25.03) | 1610 (25.37) | 165 (22.09) |  |
| Annual household income, n (%) |  |  |  | < 0.001 |
| Less than \$20,000 | 963 (13.58) | 771 (12.15) | 192 (25.70) | |
| \$20,000 to 49,999 | 2214 (31.22) | 1985 (31.28) | 229 (30.66) | |
| \$50,000 to 79,999 | 1802 (25.41) | 1629 (25.67) | 173 (23.16) | |
| \$80,000 or above | 2113 (29.79) | 1960 (30.89) | 153 (20.48) | |
| BMI, n (%) |  |  |  | < 0.001 |
| <30 kg/m<sup>2</sup> | 2497 (35.21) | 2144 (33.79) | 353 (47.26) |  |
| ≥30 kg/m<sup>2</sup> | 4595 (64.79) | 4201 (66.21) | 394 (52.74) |  |
| Smoking status, n (%) |  |  |  | < 0.001 |
| Never smoke | 4255 (60.00) | 3850 (60.68) | 405 (54.22) |  |
| Former smoker | 1966 (27.72) | 1748 (27.55) | 218 (29.18) |  |
| Current smoker | 871 (12.28) | 747 (11.77) | 124 (16.60) |  |
| **Census tract-level SVI percentile ranking** |  |  |  |  |
| Overall SVI, Mean (SD) | 0.44 (0.28) | 0.44 (0.28) | 0.48 (0.29) | < 0.001 |
| Socioeconomic theme | 0.40 (0.28) | 0.40 (0.28) | 0.45 (0.29) | < 0.001 |
| Household composition theme | 0.43 (0.29) | 0.43 (0.29) | 0.46 (0.29) | 0.001 |
| Minority status & language theme | 0.55 (0.27) | 0.55 (0.26) | 0.56 (0.27) | 0.147 |
| Housing/transportation theme | 0.48 (0.28) | 0.48 (0.28) | 0.51 (0.28) | < 0.001 |

Abbreviations: BMI, body mass index; SVI, Social Vulnerability index.
Data were presented as mean ± SD or n (%).
P value from chis-square test or t-test.

### Distribution and correlation of GIS-based hazards by SVI

**Table 1** shows that organic matter is the most abundant chemical constituents in fine particulate matter (PM_2.5_), while toluene has the highest average exposure level among VOCs of TRI emissions. Hazardous waste site was located closest to participants’ residences relative to other toxicant sites. **Figure 1 (A)** illustrates that air pollutants, VOC emissions, highway density, and proximity to most of the hazardous sites were positively correlated with each other. **Figure 1 (B)** indicates that hazards in the high SVI group showed stronger pairwise correlations (*r* ≥ 0.5) compared to the low SVI group. Exposure distribution by overall SVI was further presented in **Figure S1**.

**Figure 1.**
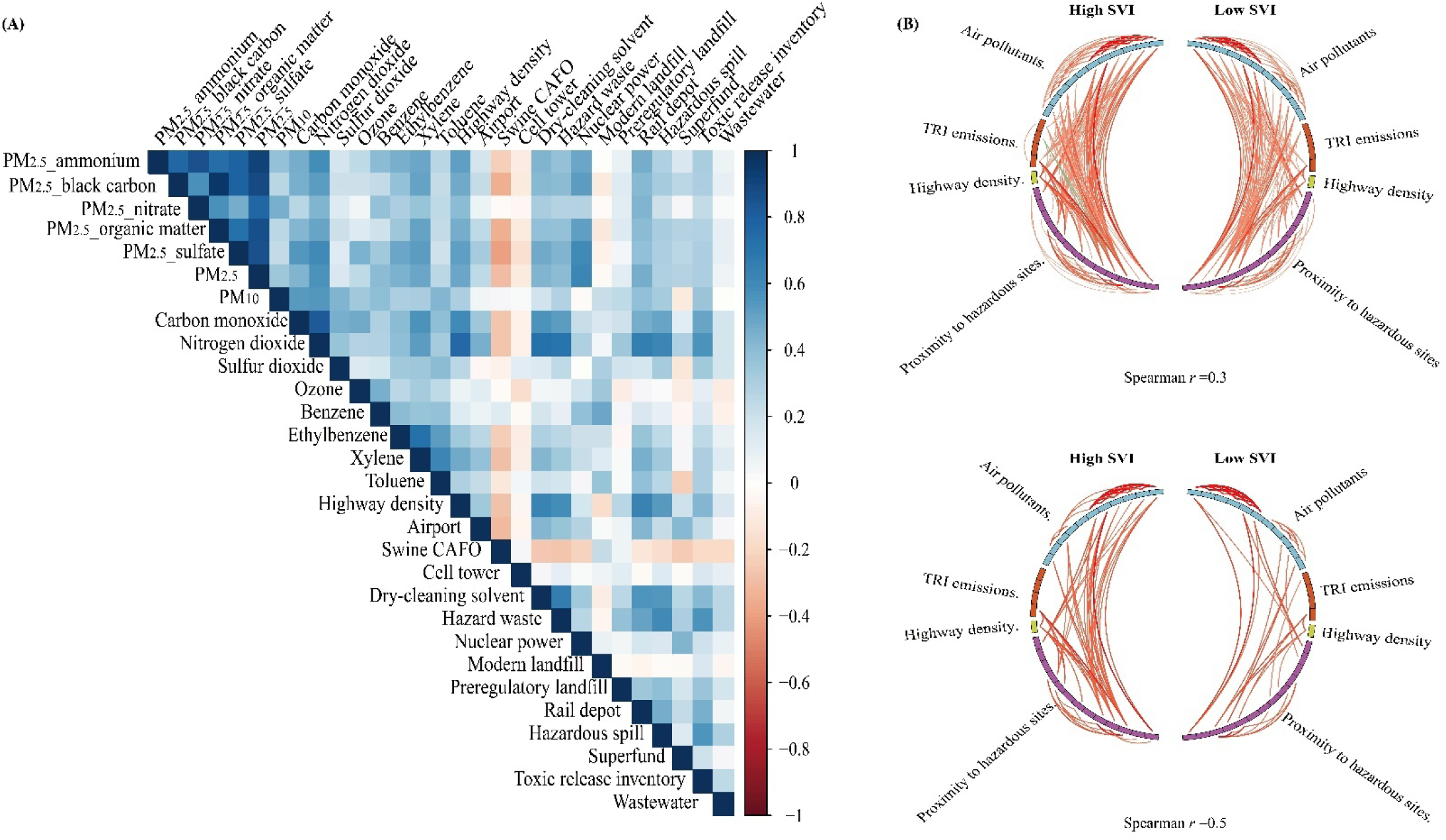
(A) Pairwise correlation of neighborhood hazards in the total population and (B) exposome globe in the high and low SVI groups (N=7092) Log_2_-transformed distance to hazard sources was inversely coded to indicate the proximity (new value= max value of the variable - original value), with higher value indicating closer to the hazard sources. (A) Spearman *r* was shown in the heatmap, with dark blue blocks indicating positive correlation near 1 and dark red blocks indicating negative correlation near -1. (B) Exposome correlation globe showing the correlations of hazards in the high (left-half) and low SVI groups (right-half). Spearman *r* greater than 0.3 or 0.5 were shown as connections in the globes, respectively. Red line denotes positive correlation, and dark green line denotes a negative one. Color intensity and line width are proportional to the size of the correlation. Within-class and between-class correlations are shown outside and inside of the track, respectively.

### Association between residential hazard exposures and asthma by SVI

Figure 2 **(A)** illustrates that proximity to the nearest dry-cleaning solvent contamination site was associated with asthma in the total population (OR=1.08, 95% CI: 1.04,1.13; FDR= 0.010) with adjustment for covariates. When stratified by SVI [Figure 2 **(B)-(C)]**, eight environmental hazards were positively associated with asthma in the high SVI group (FDR< 0.05), with organic matters of PM_2.5_ being the top hazard (OR=1.14, 95% CI: 1.06,1.23 per IQR exposure increase; FDR= 0.006) followed by the proximity to dry-cleaning solvent contamination site and black carbon in PM_2.5_. No significant associations were observed in the low SVI group. Most hazards exposures showed linear relationship with asthma (**Figures S2-S3**)

**Figure 2.**
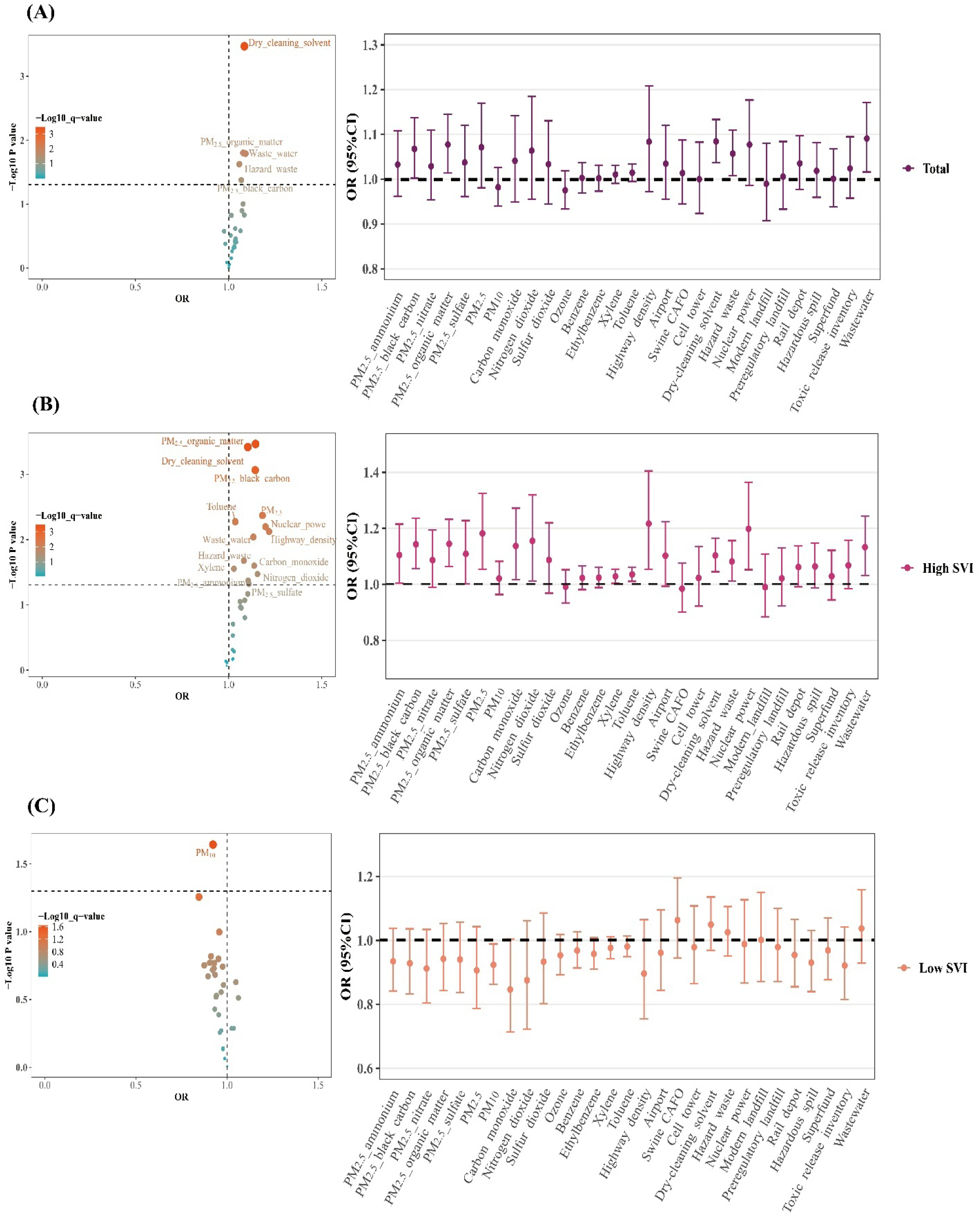
Exposome-wide association of neighborhood hazard exposures with asthma in the whole population and the subgroups by census tract-level SVI (n=7092) (A) Volcano and forest plots of the coefficient estimates and *P* value in the ExWAS analysis in the whole population. (B) Volcano and forest plots of the coefficient estimates and *P* value in the ExWAS analysis in high SVI group (C) Volcano and forest plots of the coefficient estimates and *P* value in the ExWAS analysis in the low SVI group The dashed horizontal line represents *P* =0.05 in the volcano plot; The dotted horizontal line represents *OR*=1 in the forest plot. The exposures with the *P* value<0.05 are labeled. Abbreviations: ExWAS= exposome-wide association study; OR =odds ratio. The ExWAS analysis was adjusted for age, sex, race, highest education, annual household income, and BMI. The logistic model was adjusted for age, sex, race, employment, highest education, annual household income, BMI, and smoking status. Variables of distance to hazardous sites were inversely coded to indicate the proximity to the hazardous sites (new value= max value of the variable - original value) in the regression model, with higher value indicating closer to the hazard sources. [10 exposures (most of them are air pollutants) with significant difference in their association with asthma between the high and low overall SVI groups can be further labeled or marked, eg, * *P* _interaction_< 0.05; ** *P* _interaction_ < 0.01

For 14 hazards with FDR< 0.10 in the association analysis in the high SVI group (**Table S2**), their interactions with specific SVI theme were examined. Figure 3 and **Table S3** show increased asthma odds was primarily associated with toluene, and PM_2.5_ constituents, and nuclear power station in neighborhoods with high socioeconomic and household composition vulnerability, but not in low-vulnerability neighborhoods (*P* _interaction_<0.05; *FDR* _interaction_ <0.10). Additionally, highway density, carbon monoxide (CO), and PM_2.5_ and its constituents showed strengthened positive association with asthma for neighborhoods with high socioeconomic or housing/transportation vulnerability (*P* _interaction_<0.05; *FDR* _interaction_ <0.10).

**Figure 3.**
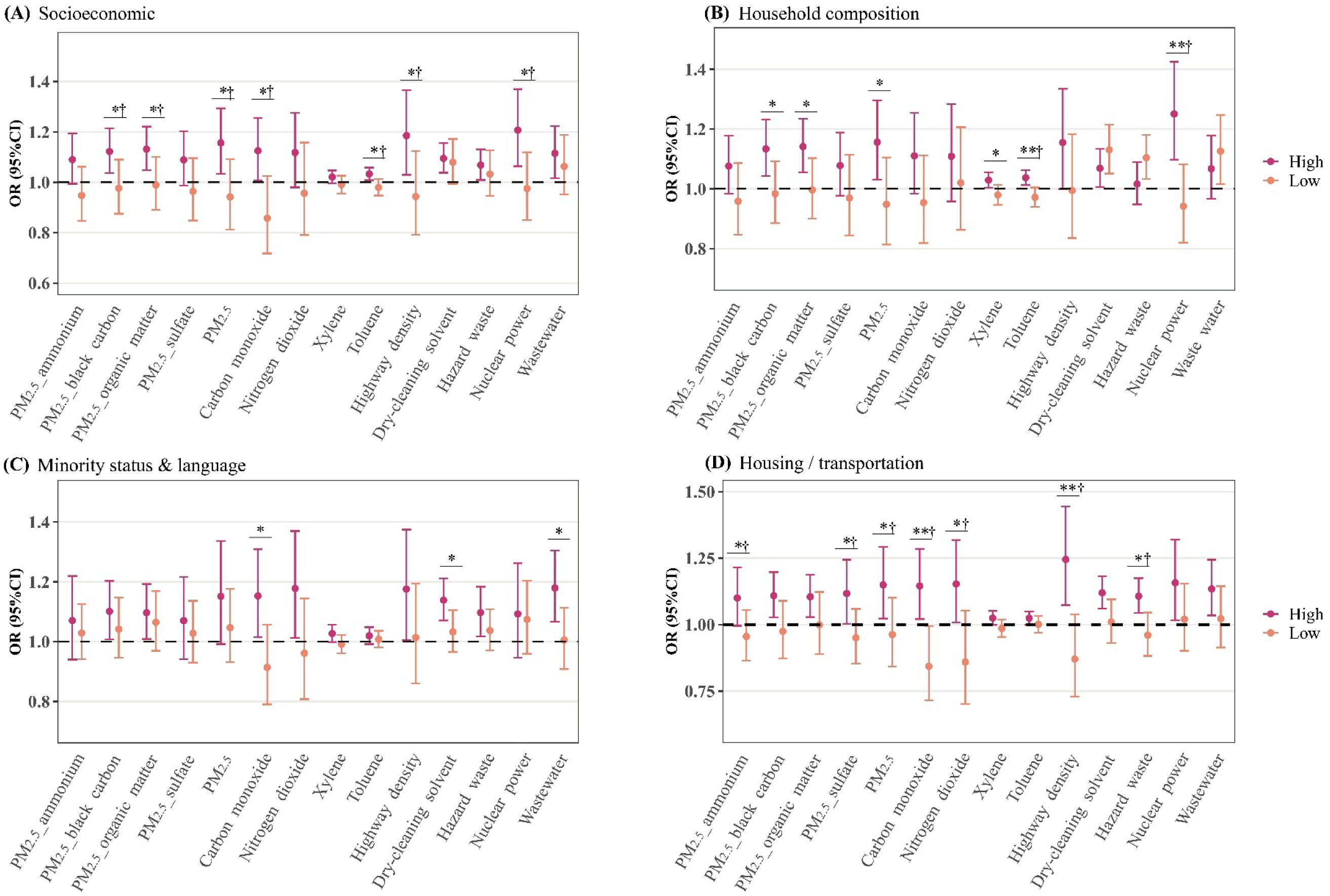
The effect modification of specific themes of SVI on the association between neighborhood hazards on adult asthma (N=7092) Exposures with FDR<0.10 (n=14) in the high SVI group were further selected to examine their interaction with four SVI themes The logistic model was adjusted for age, sex, race, employment, highest education, annual household income, BMI, and smoking status. Variables of distance to hazardous sites were inversely coded to indicate the proximity to the hazardous sites (new value= max value of the variable - original value) in the regression model, with higher value indicating closer to the hazard sources. [the exposures (most of them are air pollutants) with significant difference in their association with asthma between the high and low SVI-theme groups can be further labeled or marked in the plots] \**P* _interaction_< 0.05; ** *P* _interaction_ < 0.01; † *FDR* _interaction_ < 0.10

### Mixture exposure to hazards and asthma by SVI

Figure 4 illustrates that the joint effect of all hazards on asthma was significant only in the high SVI group. Specially, a quintile increase in exposures was associated with a 35% (95%CI: 6%, 73%) higher odds of asthma in the high SVI group, which was stronger than it in the low SVI group (*P* _for difference_ =0.038). Toluene, proximity to the nearest dry-cleaning sites, organic matter in PM_2.5,_ and CO contributed over 10% to the overall mixture effect (weight index: 0.110-0.154). Other consistent positive contributors across SVI groups include proximity to wastewater treatment plants and nuclear power stations. High socioeconomic, minority & language, and housing/transportation-related vulnerability also modify mixture effects of hazards (*P* _for difference_ **=** 0.079-0.092, **Figure S4**)

**Figure 4.**
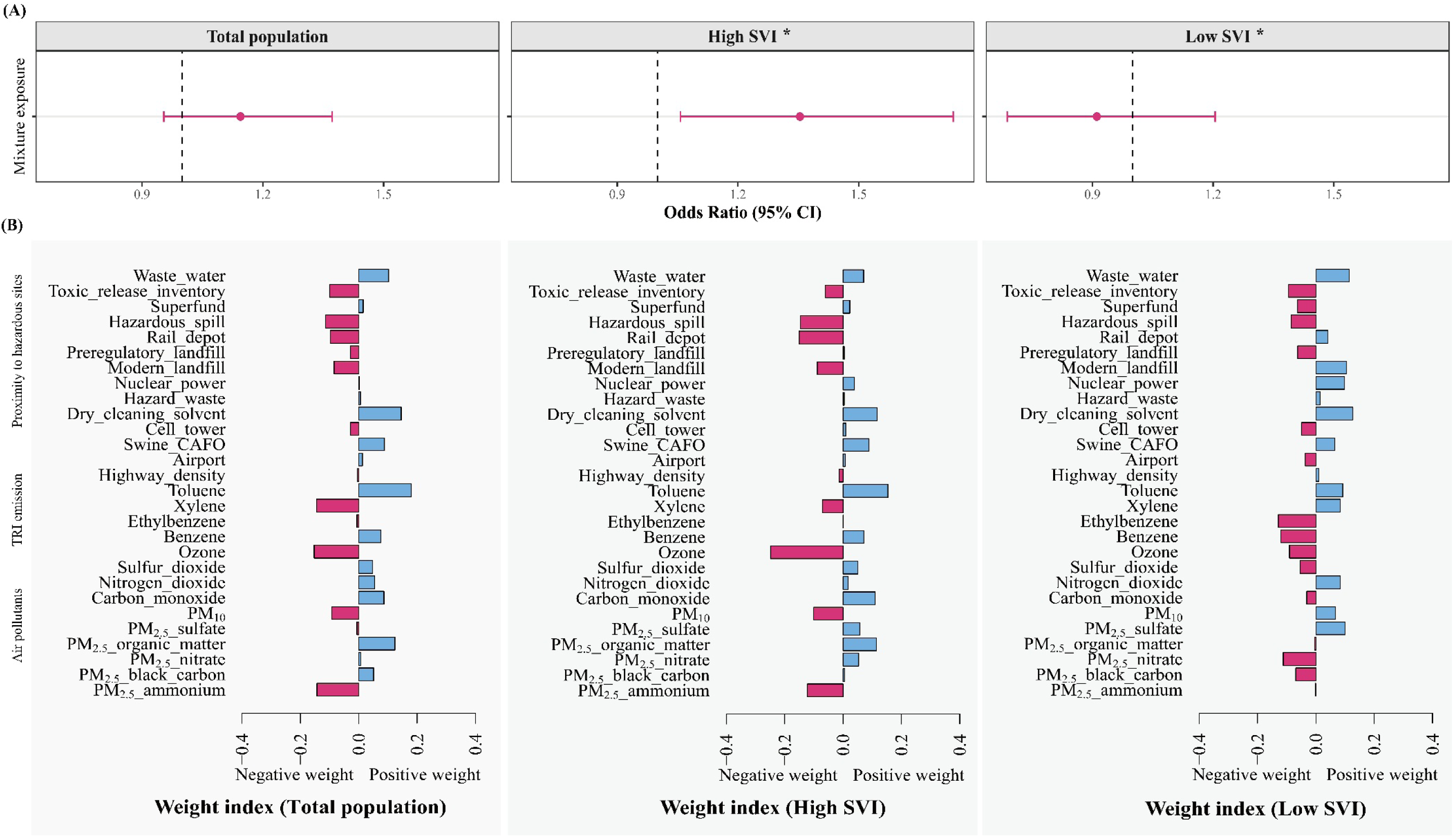
(A) The joint effects and (B) weight indexes of mixture exposure to neighborhood hazards and adult asthma in total population and the high and low SVI groups * *P* _for difference_ =0.038; The difference in mixture effects of hazards in the high SVI and low SVI groups was tested by a two-sample z-test of their estimates and standard errors. Estimated effects associated with one quintile increase in exposures to hazardous mixture in the quantile g-computation model. The model was adjusted for age, sex, race, employment, highest education, annual household income, BMI, and smoking status.

### Sensitivity analysis

When using 5-km buffer of TRI emission variables and highway density, the positive association between toluene and asthma were consistent in the high SVI group (**Figure S5**). When additionally adjusted for parental asthma, most observed positive associations with asthma were similar in the high SVI group. (**Figure S6**).

## Discussion

This study identified proximity to contaminated dry-cleaning sites as the predominant neighborhood hazard for asthma in the overall population. In the high SVI group, seven additional risk factors were identified and exposure to hazard mixture was significantly associated with asthma, with toluene, dry-cleaning solvent contaminated sites, organic matter in PM_2.5_, and CO being primary contributors. Socioeconomic and housing/transportation-related vulnerabilities interacted with these key hazards and mixture exposure on asthma.

Residential proximity to the nearest sites with dry-cleaning contamination was consistently demonstrated a positive association with adult asthma in both single-exposure and mixture analyses across SVI groups. This association may be explained by vapor intrusion of chlorinated VOCs in the contaminated sites, which can migrate from contaminated soil or underground water into indoor air. Tetrachloroethylene (PERC) – one of the most common dry-cleaning solvents – can degrade into trichloroethylene (TCE) in the environment. The adverse effects of TCE on the immune system have been well established, and TCE-induced autoimmune and vascular diseases may impair pulmonary function. ^29^ Although the lungs are not major target organ, studies have reported associations between TCE and respiratory tract disorders including asthma and chronic bronchitis. ^30^ ^31^ A Tunisian study found that PERC and TCE levels in the dry-cleaning facilities located within residential settings were at high risk of inducing immune effects, potentially threatening surrounding communities. ^32^ Similarly, a study in Paris dwellings reported elevated domestic PERC levels within 250 m of dry-cleaning facilities. ^33^ In our study, the dose-response curve indicated that asthma risk increased more rapidly within 900 m of sites. However, further investigations are warranted to confirm these findings and elucidate the specific toxicants responsible for heightened asthma risk.

In the high SVI group, more specific hazardous exposures were identified, revealing co-exposure patterns and a significant joint association with asthma. Residents of disadvantaged neighborhoods may experience multi-source pollution and worse cumulative health impacts. Positive contributors to mixture effects on asthma included VOC emissions, air pollutants, and several pollution sources. These key hazards likely share common origins (e.g., engine exhaust) ^34,35^ and biological pathways (e.g., oxidative stress or immune-mediated inflammation) ^36–38^, partially explaining their correlations and combined effects on asthma. Notably, toluene – a VOC species contributing to secondary organic aerosol formation – emerged as a key contributor to the mixture effect. ^34^, Our study quantified toluene emissions using the TRI data, which tracks toxic chemical releases from industrial and federal facilities, supporting previous studies linking toluene to asthma risk. ^38–42^ For example, a time-series study in Taiwan associated ambient toluene levels with asthma hospitalizations ^42^, while a South Korean study found industrial toluene contamination correlated with higher respiratory disease prevalence. ^39^ Measuring urinary metabolites, a Chinese case-control study found co-exposure to toluene, benzene, and polycyclic aromatic hydrocarbons (PAH), was also associated with increased DNA and lipid oxidative damages and airway inflammation in asthmatic children. ^38^

For air pollutants, systematic reviews have summarized a positive association between PM_2.5_ and asthma ^43,44^, while few studies have decomposed the specific toxicities of its constituents. ^45–47^ Consistent with cohort studies on childhood asthma ^45,46^, we found organic matters, black carbon, and sulfate individually and jointly contributed to adult asthma risk in high-SVI populations, probably due to their high oxidative potentials to induce airway inflammation and hyperresponsiveness. ^48–50^ Organic matter comprises the largest fraction of PM_2.5_ and showed greater contribution than other constituents. PAHs, as major organic compounds in PM_2.5_, have been shown to promote TH_17_/Treg imbalance and aggravate asthma through AhR-dependent pathways. ^37^ In addition to PM_2.5_, CO and NO_2_ were associated with decreased lung function and adult asthma, although exposure levels between studies are an important source of heterogeneity ^43,51–53^. Despite recent declines in air pollution levels in the U.S. (mean levels of 10.6 μg/m^3^ for PM_2.5_, 0.35 μg/m^3^ for CO, and 12.0 μg/m^3^ for NO_2_ in this study), health benefits remain unequally distributed. ^14^ Prior research in North Carolina observed neighborhoods with higher poverty, lower education, and greater neighborhood deprivation and a higher proportion minority had higher levels of air pollutants ^54^. These findings underscore ongoing efforts are needed to mitigate air-pollution related asthma risk for socially disadvantaged populations.

The vulnerability to hazard-related asthma risks was primarily driven by socioeconomic and housing/transportation factors in this study. Poor socioeconomic status, such as poverty, unemployment, and low education, can predispose individuals to be more likely to be harmed to specific hazards. ^2^ Housing conditions are shaped by social forces and consist of physical components (e.g., location, density, and sanitation), mediating exposure to toxicants in residential environment. ^11^ Neighborhood-level poor housing quality indicated by housing structure and type may translate social adversities into population health disparities, including asthma prevalence. ^11,55^ Previous research have documented domain-specific SVI-related asthma risk ^12,56–58^, including a U.S. metropolitan study linking neighborhood-level vulnerability in demographic, economic, and residential (including housing structures) domains to greater pediatric asthma morbidity. ^12^ Studies examining social-environmental interactions further showed stronger pollution-asthma associations in neighborhoods with higher deprivation levels ^16^ and lower socioeconomic status ^17^ measured by education, income, poverty, unemployment, household type, and transportation. High SVI neighborhoods are often located closer to industrial facilities and major roadways. In these disinvested communities with degraded environments, loss of natural resources and limited access to quality healthcare and other essential services can exacerbate health risk inequities. ^14^ Our findings align with previous evidence, emphasizing on prioritizing public health resources for vulnerable neighborhoods in key domains.

Our study effectively utilizes GIS data and a mixture approach to map the relationships between various neighborhood toxicants and asthma. Although built environment has been previously explored for asthma ^8–10^, residential proximity to multiple neighborhood pollution sources is rarely evaluated separately or collectively with other hazards. Our research demonstrated their co-exposure effects and interaction with SVI, providing implications for census-tract interventions. However, several limitations were noted. First, although geospatial estimates offer insight into proximity to hazardous sources, they cannot directly assess toxicants levels as personal monitoring methods do. For VOC emissions, exposure levels were inferred at predefined buffers, while the optimal radius reflecting affected areas is unclear. Nonetheless, we prioritized a 10-km buffer to capture larger exposure variations, and our key finding on toluene was consistent with a 5-km buffer in the sensitivity analysis. Second, we averaged annual concentrations of air pollutants during study periods, based on the assumption that long-term air pollutants have cumulative impacts on the development of chronic communicable diseases like asthma. However, short-term exposures may be related to severe asthma exacerbations or mortality ^59,60^, which could not be assessed in this cross-sectional study and need further investigations. Third, this study relied on publicly available exposure datasets from federal or state environmental protection agencies. While this enables reproducibility, it also restricts the range of pollutants analyzed. Fourth, this study focused on neighborhood hazards and social vulnerability which need public health interventions and regulations; however, individual-level modifiable behavioral factors and household environment were not included. Future exposome study should consider integrating diverse datasets from multiple disciplines over the life course to more comprehensively understand asthmas etiology.

## Conclusions

This North Carolina population-based study explored various neighborhood hazards, linking proximity to pollutant sources to increased asthma risk, especially dry-cleaning solvent contamination. Despite generally low air pollutants concentrations, toluene, organic matter in PM_2.5_, and CO continued to pose asthma risk to socially vulnerable neighborhoods. Moreover, neighborhoods with high socioeconomic status and housing / transportation -specific vulnerability were more likely to experience multi-hazard exposures and related asthma risk. Our findings highlighted the importance of targeted control measures for emission sources of key hazards and community-level interventions to address health disparities caused by social and environmental factors.

## Data Availability

The data will be available under controlled access to comply with polices and process of the PEGS data and meet applicable NIH requirements for data sharing and that the privacy and confidentiality of human subjects are protected. Researchers seeking access to PEGS data may contact the PEGS Executive Leadership Committee with details from the PEGS websites

