## Supplementary material for "Geospatial-based neighborhood hazards and asthma risk in adults: interaction with social vulnerability": Table S1

Supplementary materials

**Table S1 Description of CDC/ATSDR social vulnerability index**

| SVI themes | Components |
| --- | --- |
| Socioeconomic | Person below poverty |
|  | Persons age 16+ unemployed |
|  | Per-capita income |
|  | Persons age 25+ with no high school diploma |
| Household composition | Person age 65 and older |
|  | Person age 17 and younger |
|  | Civilian with a disability |
|  | Single parent households |
| Minority Status & Language | Proportion minority (persons except white, non-Hispanic) |
|  | English proficiency (persons age 5+ who speak Engligh less than well) |
| Housing/transportation | Multi-unit structures |
|  | Mobile homes (households living in mobile homes) |
|  | Crowding (households with more people than rooms) |
|  | No vehicle (households with no vehicle) |
|  | Group quarters (persons living in institutionalized group quarters [e.g. nursing homes, military bases]) |

Table S2 The interaction of neighborhood hazards with overall SVI on adult asthma

|  |  | Overall low SVI | Overall high SVI |  |  |
| --- | --- | --- | --- | --- | --- |
| Exposure |  | OR (95% CI) | OR (95% CI) | *P* _interaction_ | *FDR* _interaction_ |
| Air pollutants | PM_2.5-_ammonium | 0.93 (0.84, 1.04) | **1.10 (1.00, 1.22)** | **0.017** | **0.046** |
|  | PM_2.5__black carbon | 0.93 (0.83, 1.04) | **1.14 (1.06, 1.24)** | **0.032** | **0.049** |
|  | PM_2.5__organic matter | 0.94 (0.84, 1.05) | 1.14 (1.06, 1.23) | 0.068 | 0.095 |
|  | PM_2.5__sulfate | 0.94 (0.84, 1.06) | **1.11 (1.00, 1.23)** | **0.030** | **0.049** |
|  | PM_2.5_ | 0.91 (0.79, 1.04) | **1.18 (1.05, 1.33)** | **0.010** | **0.046** |
|  | Carbon monoxide | 0.85 (0.71, 1.00) | **1.14 (1.02, 1.27)** | **0.018** | **0.046** |
|  | Nitrogen dioxide | 0.88 (0.72, 1.06) | 1.15 (1.01, 1.32) | 0.124 | 0.158 |
| TRI emissions | Xylene | 0.98 (0.94, 1.01) | **1.03 (1.00, 1.05)** | **0.016** | **0.046** |
|  | Toluene | 0.98 (0.95, 1.01) | **1.04 (1.01, 1.06)** | **0.020** | **0.046** |
| Highway density | Highway density | 0.90 (0.75, 1.06) | **1.22 (1.05, 1.40)** | **0.013** | **0.046** |
| Proximity to hazardous sites | Dry-cleaning solvent | 1.05 (0.97, 1.14) | 1.10 (1.04, 1.16) | 0.786 | 0.786 |
|  | Hazard waste | 1.03 (0.95, 1.11) | 1.08 (1.01, 1.16) | 0.631 | 0.679 |
|  | Nuclear power | 0.99 (0.87, 1.13) | **1.20 (1.05, 1.36)** | **0.029** | **0.049** |
|  | Wastewater | 1.04 (0.93, 1.16) | 1.13 (1.03, 1.24) | 0.517 | 0.603 |

The model was adjusted for age, sex, race, employment, highest education, annual household income, BMI, and smoking status.

Table S3 The interaction of neighborhood hazards with specific SVI themes on adult asthma

|  |  | Low SVI | High SVI |  |  |
| --- | --- | --- | --- | --- | --- |
| SVI theme / Exposure |  | OR (95% CI) | OR (95% CI) | *P* _interaction_ | *FDR* _interaction_ |
| **Socioeconomic** | |  |  |  |  |
| Air pollutants | PM_2.5__ammonium | 0.95 (0.85, 1.06) | 1.09 (0.99, 1.19) | 0.057 | 0.099 |
|  | PM_2.5__black carbon | 0.98 (0.88, 1.09) | **1.12 (1.04, 1.21)** | **0.046** | **0.099** |
|  | PM_2.5__organic matter | 0.99 (0.89, 1.10) | **1.13 (1.05, 1.22)** | **0.049** | **0.099** |
|  | PM_2.5__sulfate | 0.96 (0.85, 1.10) | 1.09 (0.99, 1.20) | 0.122 | 0.190 |
|  | PM_2.5_ | 0.94 (0.81, 1.09) | **1.16 (1.03, 1.29)** | **0.029** | **0.099** |
|  | Carbon monoxide | 0.86 (0.72, 1.03) | **1.13 (1.01, 1.26)** | **0.012** | **0.085** |
|  | Nitrogen dioxide | 0.96 (0.79, 1.16) | 1.12 (0.98, 1.28) | 0.210 | 0.267 |
| TRI emissions | Xylene | 0.99 (0.96, 1.03) | 1.02 (1.00, 1.05) | 0.179 | 0.250 |
|  | Toluene | 0.98 (0.95, 1.01) | **1.03 (1.01, 1.06)** | **0.011** | **0.085** |
| Highway density | Highway density | 0.94 (0.79, 1.12) | 1.19 (1.03, 1.37) | 0.048 | 0.099 |
| Proximity to hazardous sites | Dry-cleaning solvent | 1.08 (0.99, 1.17) | 1.10 (1.04, 1.16) | 0.757 | 0.757 |
|  | Hazard waste | 1.03 (0.95, 1.13) | 1.07 (1.01, 1.13) | 0.569 | 0.613 |
|  | Nuclear power | 0.98 (0.85, 1.12) | **1.21 (1.06, 1.37)** | **0.020** | **0.092** |
|  | Wastewater | 1.06 (0.95, 1.19) | 1.12 (1.02, 1.22) | 0.514 | 0.600 |
| **Household composition** | |  |  |  |  |
| Air pollutants | PM_2.5__ammonium | 0.96 (0.85, 1.09) | 1.08 (0.98, 1.18) | 0.156 | 0.243 |
|  | PM_2.5__black carbon | 0.98 (0.88, 1.09) | 1.13 (1.04, 1.23) | **0.039** | 0.110 |
|  | PM_2.5__organic matter | 1.00 (0.90, 1.10) | 1.14 (1.06, 1.23) | **0.040** | 0.110 |
|  | PM_2.5__sulfate | 0.97 (0.84, 1.11) | 1.08 (0.98, 1.19) | 0.246 | 0.294 |
|  | PM_2.5_ | 0.95 (0.81, 1.10) | 1.16 (1.03, 1.30) | **0.047** | 0.110 |
|  | Carbon monoxide | 0.95 (0.82, 1.11) | 1.11 (0.98, 1.25) | 0.138 | 0.242 |
|  | Nitrogen dioxide | 1.02 (0.86, 1.21) | 1.11 (0.96, 1.28) | 0.512 | 0.512 |
| TRI emissions | Xylene | 0.98 (0.95, 1.01) | 1.03 (1.00, 1.05) | **0.024** | 0.110 |
|  | Toluene | 0.97 (0.94, 1.00) | **1.04 (1.01, 1.06)** | **0.002** | **0.022** |
| Highway density | Highway density | 0.99 (0.84, 1.18) | 1.15 (1.00, 1.33) | 0.192 | 0.269 |
| Proximity to hazardous sites | Dry-cleaning solvent | 1.13 (1.05, 1.21) | 1.07 (1.01, 1.13) | 0.252 | 0.294 |
|  | Hazard waste | 1.10 (1.03, 1.18) | 1.02 (0.95, 1.09) | 0.085 | 0.170 |
|  | Nuclear power | 0.94 (0.82, 1.08) | **1.25 (1.10, 1.42)** | **0.003** | **0.024** |
|  | Wastewater | 1.13 (1.02, 1.25) | 1.07 (0.97, 1.18) | 0.497 | 0.512 |
| **Minority Status & Language** | |  |  |  |  |
| Air pollutants | PM_2.5__ammonium | 1.03 (0.94, 1.12) | 1.07 (0.94, 1.22) | 0.489 | 0.587 |
|  | PM_2.5__black carbon | 1.04 (0.95, 1.15) | 1.10 (1.01, 1.20) | 0.299 | 0.465 |
|  | PM_2.5__organic matter | 1.06 (0.97, 1.17) | 1.10 (1.01, 1.19) | 0.494 | 0.587 |
|  | PM_2.5__sulfate | 1.03 (0.93, 1.14) | 1.07 (0.94, 1.22) | 0.550 | 0.592 |
|  | PM_2.5_ | 1.05 (0.93, 1.18) | 1.15 (0.99, 1.34) | 0.230 | 0.403 |
|  | Carbon monoxide | 0.91 (0.79, 1.06) | **1.15 (1.02, 1.31)** | **0.018** | 0.132 |
|  | Nitrogen dioxide | 0.96 (0.81, 1.14) | 1.18 (1.01, 1.37) | 0.056 | 0.196 |
| TRI emissions | Xylene | 0.99 (0.96, 1.02) | 1.03 (1.00, 1.06) | 0.073 | 0.206 |
|  | Toluene | 1.01 (0.98, 1.04) | 1.02 (0.99, 1.05) | 0.503 | 0.587 |
| Highway density | Highway density | 1.01 (0.86, 1.19) | 1.18 (1.01, 1.37) | 0.113 | 0.263 |
| Proximity to hazardous sites | Dry-cleaning solvent | 1.03 (0.96, 1.10) | **1.14 (1.07, 1.21)** | **0.028** | 0.132 |
|  | Hazard waste | 1.04 (0.97, 1.11) | 1.10 (1.02, 1.18) | 0.225 | 0.403 |
|  | Nuclear power | 1.07 (0.96, 1.20) | 1.09 (0.95, 1.26) | 0.988 | 0.988 |
|  | Wastewater | 1.01 (0.91, 1.11) | **1.18 (1.07, 1.30)** | **0.022** | 0.132 |

Continued Table S3

|  |  | Low SVI | High SVI |  |  |
| --- | --- | --- | --- | --- | --- |
| SVI theme / Exposure |  | OR (95% CI) | OR (95% CI) | *P* _interaction_ | *FDR* _interaction_ |
| **Housing/transportation** | |  |  |  |  |
| Air pollutants | PM_2.5__ammonium | 0.96 (0.87, 1.06) | **1.10 (1.00, 1.22)** | **0.047** | **0.095** |
|  | PM_2.5__black carbon | 0.97 (0.87, 1.09) | 1.11 (1.03, 1.20) | 0.065 | 0.105 |
|  | PM_2.5__organic matter | 1.00 (0.89, 1.12) | 1.10 (1.03, 1.19) | 0.159 | 0.203 |
|  | PM_2.5__sulfate | 0.95 (0.85, 1.06) | **1.12 (1.00, 1.24)** | **0.036** | **0.095** |
|  | PM_2.5_ | 0.96 (0.84, 1.10) | **1.15 (1.02, 1.29)** | **0.048** | **0.095** |
|  | Carbon monoxide | 0.84 (0.72, 0.99) | **1.15 (1.02, 1.29)** | **0.005** | **0.032** |
|  | Nitrogen dioxide | 0.86 (0.70, 1.05) | **1.15 (1.01, 1.32)** | **0.028** | **0.095** |
| TRI emissions | Xylene | 0.99 (0.95, 1.02) | 1.02 (1.00, 1.05) | 0.088 | 0.123 |
|  | Toluene | 1.00 (0.97, 1.03) | 1.02 (1.00, 1.05) | 0.275 | 0.275 |
| Highway density | Highway density | 0.87 (0.73, 1.04) | **1.25 (1.07, 1.44)** | **0.004** | **0.032** |
| Proximity to hazardous sites | Dry-cleaning solvent | 1.01 (0.93, 1.09) | 1.12 (1.06, 1.18) | 0.068 | 0.105 |
|  | Hazard waste | 0.96 (0.88, 1.05) | **1.11 (1.04, 1.17)** | **0.011** | **0.049** |
|  | Nuclear power | 1.02 (0.90, 1.15) | 1.16 (1.02, 1.32) | 0.202 | 0.218 |
|  | Waste water | 1.02 (0.91, 1.14) | 1.13 (1.03, 1.24) | 0.196 | 0.218 |

The model was adjusted for age, sex, race, employment, highest education, annual household income, BMI, and smoking status.

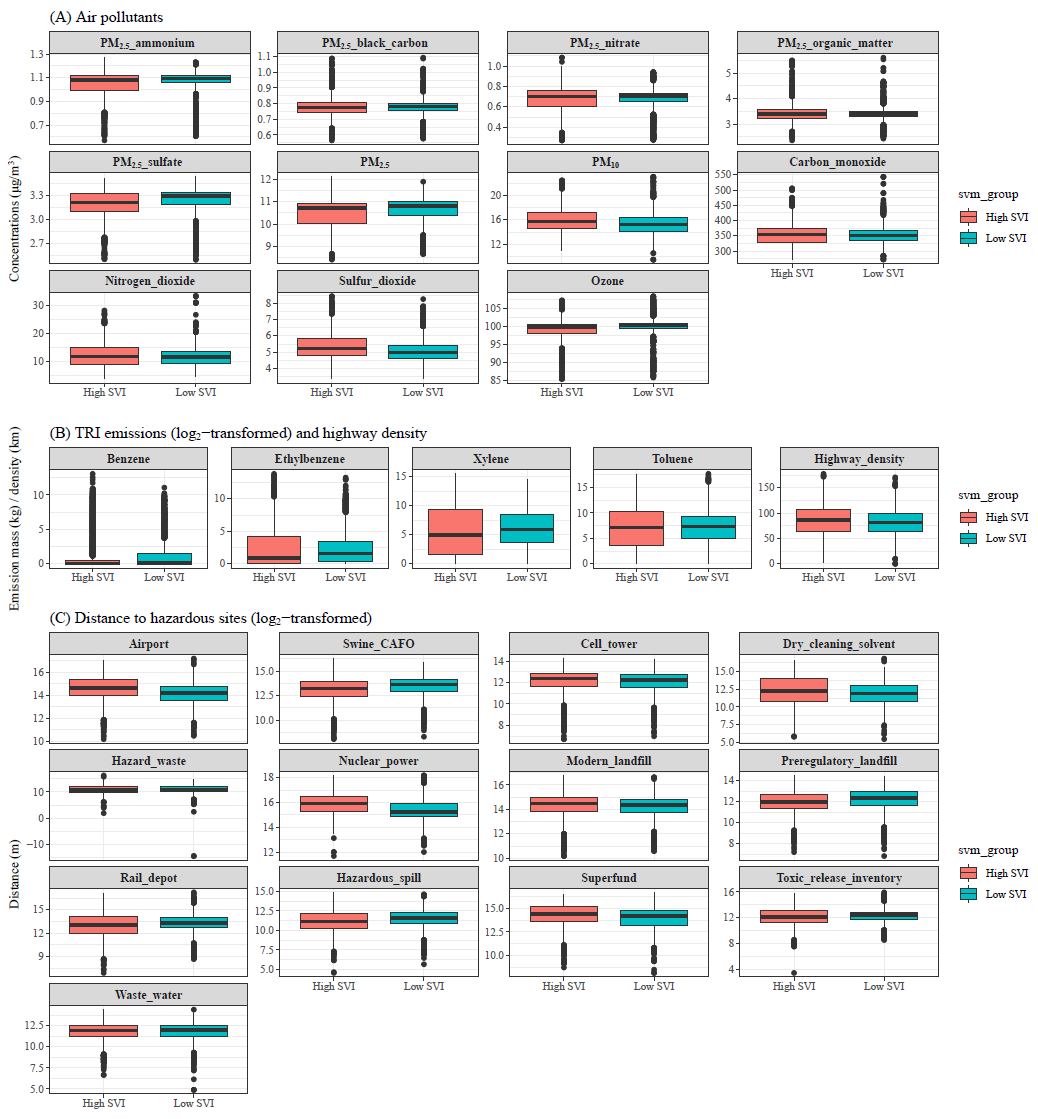
 Figure S1 The distribution of neighborhood hazards on adult asthma by the overall SVI group

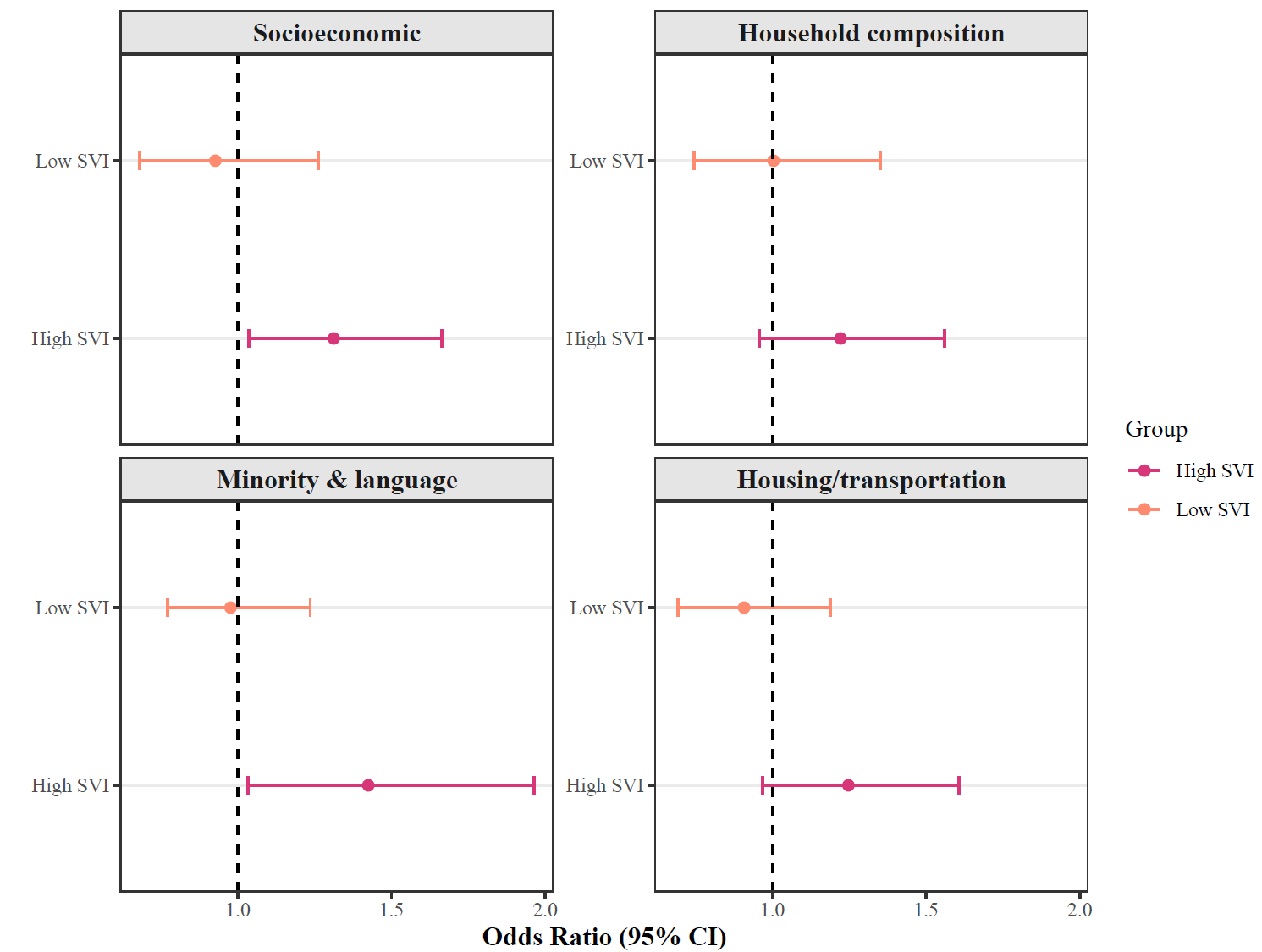
Figure S2 The joint effects of mixture exposure to neighborhood hazards on adult asthma by the specific themes of SVI groups

*P* _for difference_ **=**0.079, 0.317, 0.063, 0.092 for the difference of effects in the high and low SVI of socioeconomic, household composition, minority & language, housing / transportation themes, respectively.

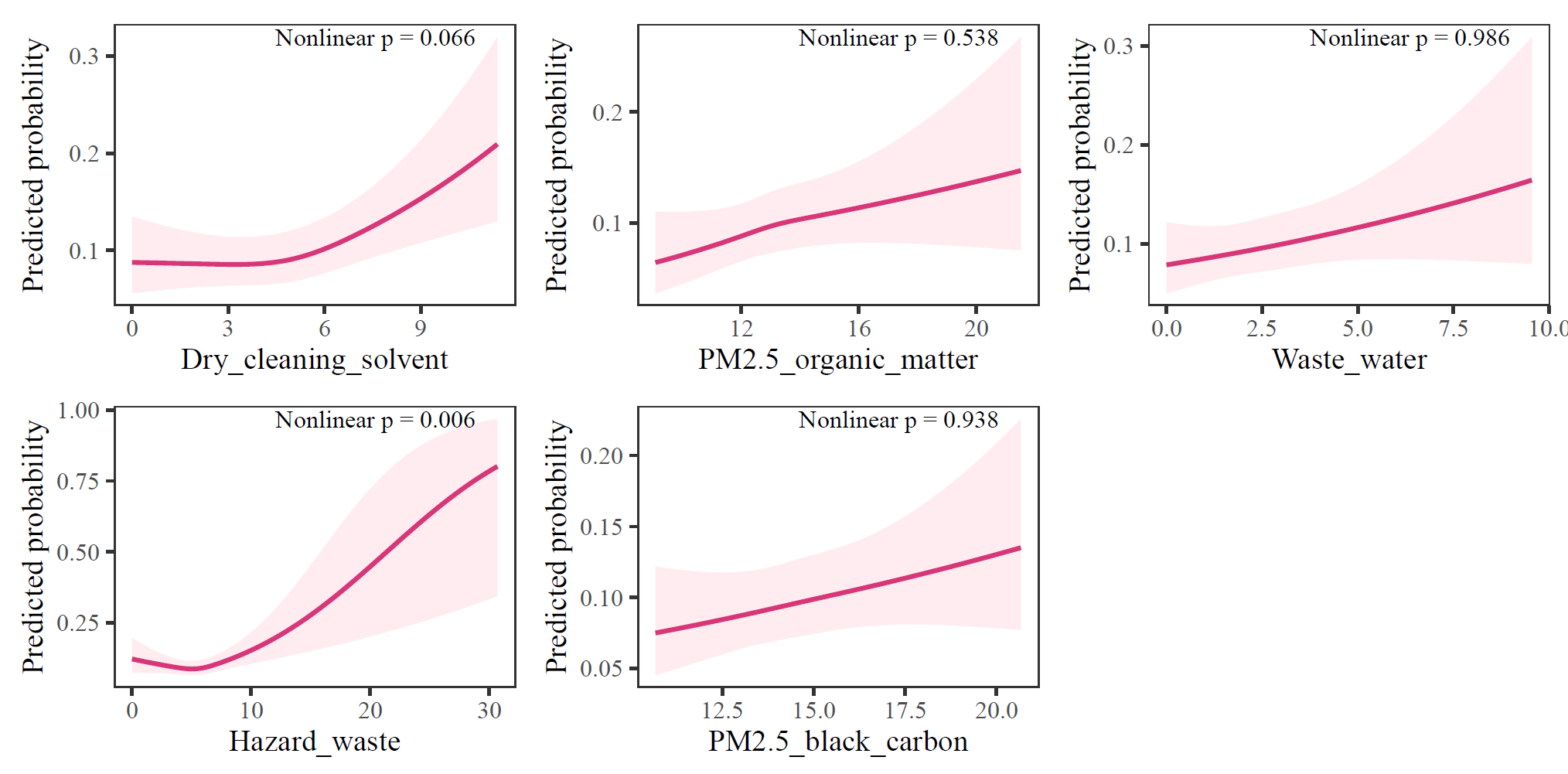

Figure S3 The dose-response probability curves of selected hazards for asthma fitted by the restricted cubic splines (knot number=3) in the total population (N=7092)

Hazards with *P*< 0.05 (FDR<0.24) in the ExWAS were tested

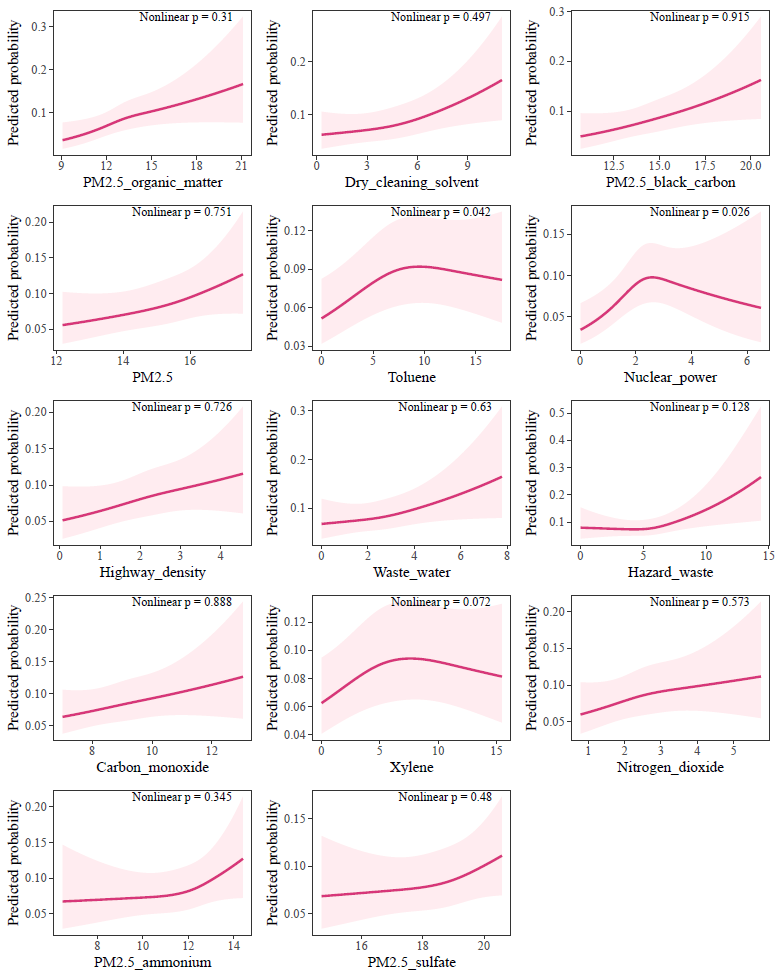
 Figure S4 The dose-response probability curves of selected hazards for asthma fitted by the restricted cubic splines (knot number=3) in the high overall SVI group(N=7092)

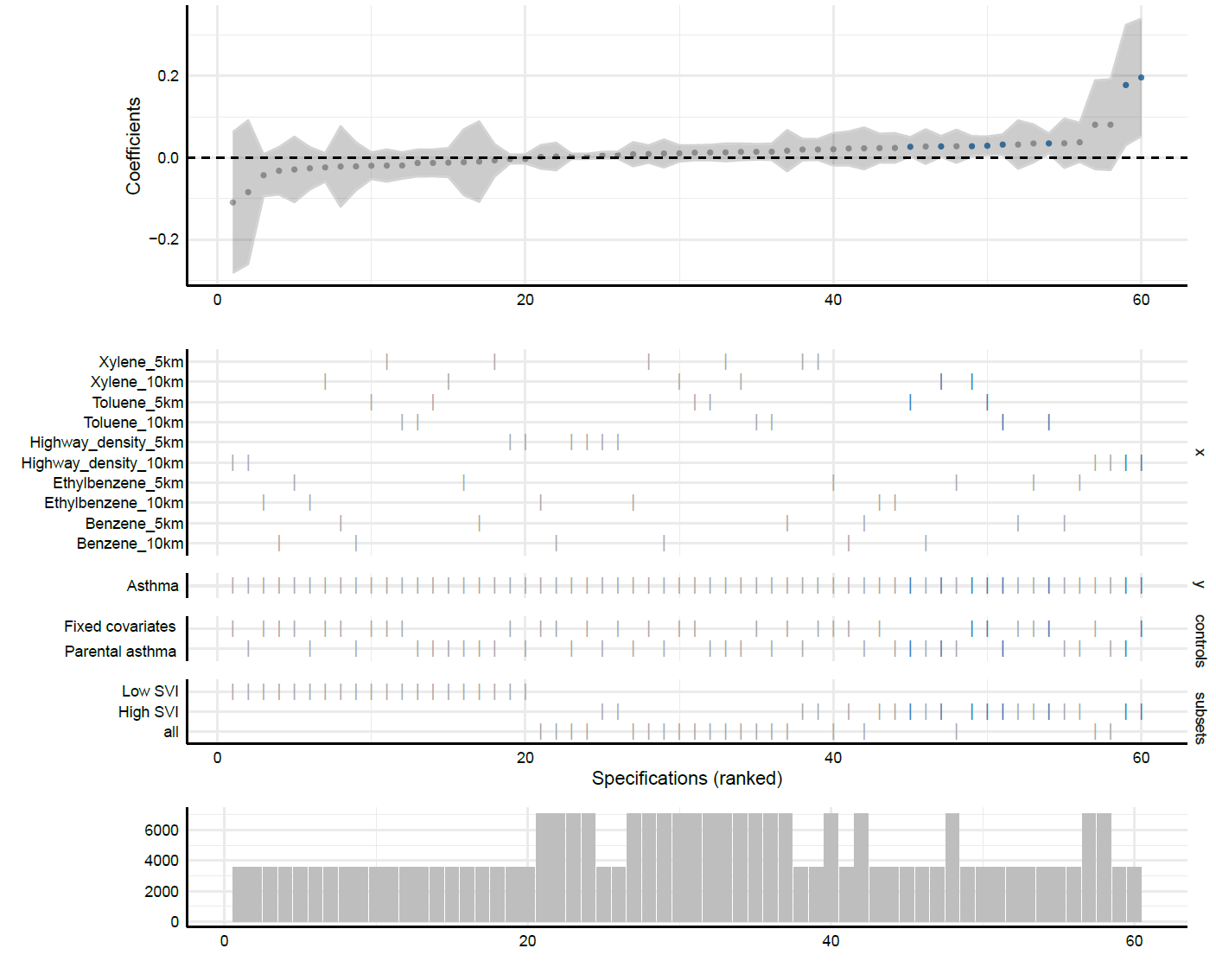
Figure S5 The specification curves for different buffers of TRI emission and highway density variables in association with asthma

Specifications in blue indicated significant estimations (*P*<0.05)

Fixed covariates: age, sex, race, employment, highest education, annual household income, BMI, and smoking status.

Results of exposures in 10-km buffer in the main analyses were also shown for comparisons.

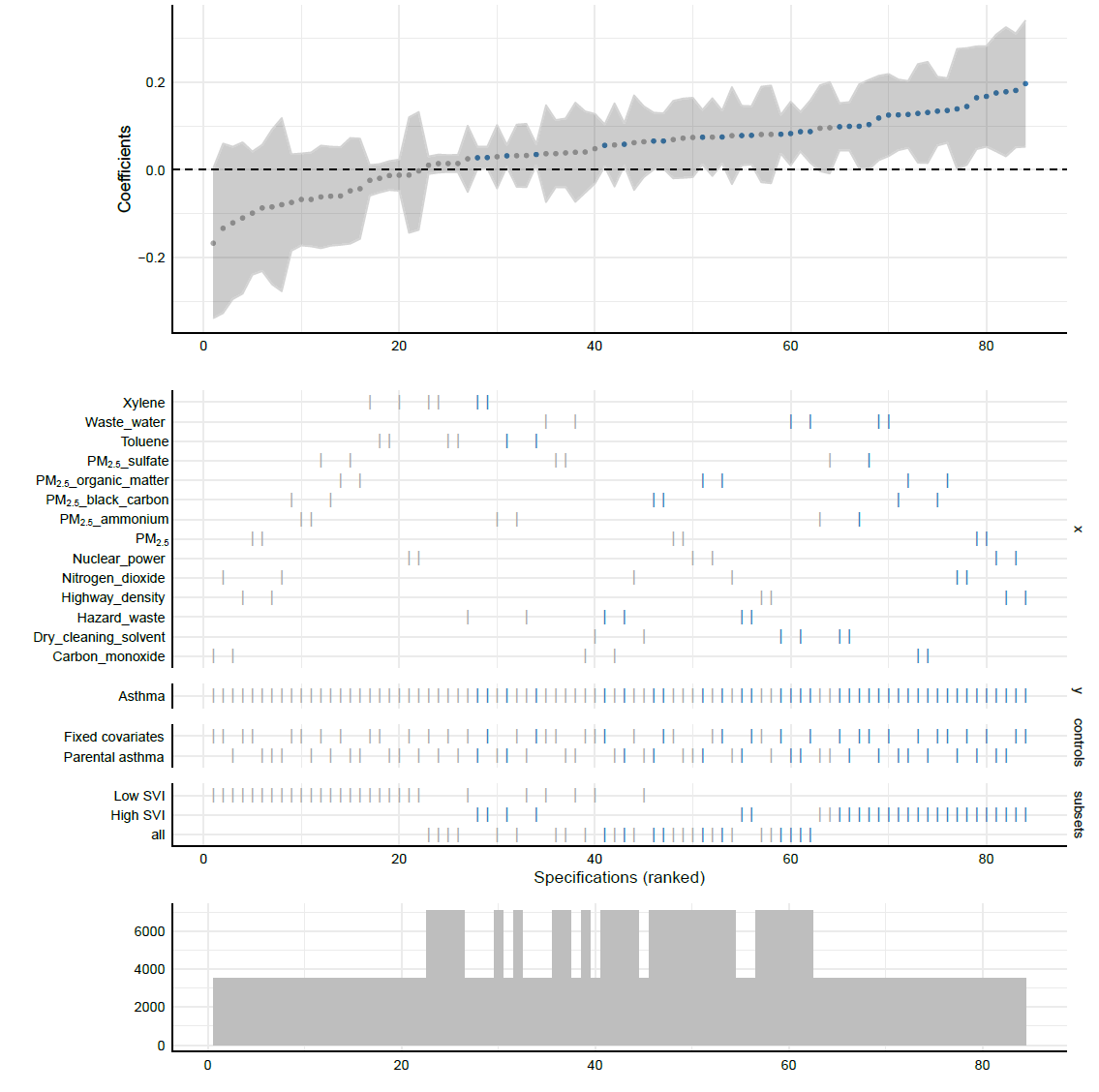
 Figure S6 The specification curves for associations between hazards and asthma with additional adjustment for parental asthma history

Exposure with FDR<0.10 in the ExWAS of main analyses in the high SVI group were included for this sensitivity analysis. Specifications in blue indicated significant estimations (*P*<0.05)

Fixed covariates: age, sex, race, employment, highest education, annual household income, BMI, and smoking status.
